# BNT162b2 mRNA vaccinations in Israel: understanding the impact and improving the vaccination policies by redefining the immunized population

**DOI:** 10.1101/2021.06.08.21258471

**Authors:** Chana Ross, Oren Spector, Meytal Avgil Tsadok, Yossi Weiss, Royi Barnea

## Abstract

By the end of February 2021, when 48% of the Israeli population was immune, the number of new positive COVID-19 cases significantly dropped across all ages. Understanding which parameters influenced this drop and how to minimize the number of hospitalizations and overall positive cases is urgently needed.

In this study we conducted an observational analysis which included COVID-19 data with over 12,000,000 PCR tests from 250 cities in Israel. In addition, we performed a simulation of different vaccination campaigns to find the optimal policy.

Our analysis revealed that cities with younger populations reached a decrease in new cases when a lower percentage of their residents were immunized, showing that median age is a crucial parameter effecting overall immunity, while other parameters appeared to be insignificant. This variance between cities is explained by recalculating the immunized population and multiplying each individual by a factor symbolizing the impact of their age on the spread on the virus. This factor is easily calculated from historical data of positive cases per age.

The simulation proves that prioritizing different age groups or changing the rate of vaccinations drastically effects the overall hospitalizations and positive cases.

**One-Sentence Summary:** understanding what influences reaching covid-19 overall immunity and how to maximize the effect of the vaccination campaign.

## INTRODUCTION

Israel launched its vaccination campaign against Coronavirus disease 2019 (COVID-19) during December 2020 by using BNT162b2 mRNA vaccination. This vaccination campaign quickly placed Israel as the country with the highest rate of vaccinated individuals per capita in the world [https://ourworldindata.org/covid-vaccinations]. From December 2020 to May 2021, over five million residents in Israel, the majority over the age of 16, received two doses of the vaccine. In practice, this means that almost 60% of the population has been immunized.

Vaccinating the population has two main goals: (1) minimizing the number of critical cases and hospitalizations^1^ and (2) containing the virus thus reducing its spread.^2,3^ As the Israeli Ministry of Health’s (MoH) vaccination policy for COVID-19 prioritized to first achieve the first goal, the vaccination campaign was aimed at first vaccinating the community at risk, including individuals older than 60, nursing home residents, healthcare workers and individuals with severe health conditions. As the campaign advanced and the percentage of vaccinated individuals in this group increased, the campaign was incrementally expanded to include younger age groups, until all individuals above 16 years were entitled for the vaccine.

During the vaccination campaign, two unique and major phases were observed in Israel: A *transition phase* - defined as the period in which the number of new COVID-19 cases dropped among vaccinated individuals, but increased among the unvaccinated, and a *community-immunity phase* where a drop in new cases was observed across all communities regardless of their vaccination status.

In-depth analysis of these two phases may assist other countries in understanding the outcome of their vaccination campaigns and improving their vaccination policies accordingly. However, clear understanding of these phases based on Israel’s macro data is challenging due to Israel’s unique demographic, culture and government interventions. Israel is known for its diversity in sectors (Ultra-orthodox Jews, Arabs and the Secular population), social economic levels, median ages and other parameters. In order to deal with those unique properties, we chose to examine the vaccination effect at a municipal level leveraging Israel’s municipal variance in culture and demographics. We conducted an observational study using daily corona demographics and economic data from 250 cities in Israel in order to understand the effect of vaccinations on the unvaccinated community and to explain the change from the *transition phase* to the *community-immunity phase*. We also attempted to understand the different optional vaccination campaign policies. This is more complex as it requires comparing different policies which were not conducted in the real world. To overcome this issue, we decided to simulate the pandemic and Israel’s unique social connectivity graph and tried to compare different vaccination policies and rates in the simulated environment.

## METHODS

### Setting

In March 2021, the MoH held a virtual data-challenge event to generate insights based on anonymized governmental data. All sectors of the Israeli research community were invited to analyze up-to-date data in three pre-determined policy related challenges. Approximately eighty participants were accepted and divided into multidisciplinary teams, comprising experts from various fields (data scientists, policy experts, clinicians and epidemiologists).

The authors’ work was selected as the winning project in the event, further enabling access to updated data following the end of the event.

### Observational study

#### Data sources

The data included two main COVID-19 tables given to us by the MoH. The first with individual identification number (random number for ethical and anonymous reasons), age group, geographic region, date of polymerase chain reaction (PCR) test for identification of COVID-19 and test result, time of hospitalization, worst case while hospitalized, COVID-19 outcome (alive/dead). The second table included vaccination data, with data about the number of vaccinations per day for different ages and geographic regions. Large portions of the aggregated data used in this work is publicly available in Hebrew divided into 6 different databases. Sensitive information, mostly related to the hospitalization status is not publicly accessible. In addition to the COVID-19 data we used data from the CBS (Central Bureau of Statistics) regarding cities median age, number of people in each age group and data about families in Israel. The COVID data included 262 cities with over 5000 citizens, but only 250 were taken into account since 12 cities did not have information on their median age Our analysis code in addition to some of the publicly available data, can be found in this Git repository https://github.com/CoronaPolicy/prediction_model.

#### Statistical analysis

In order to compare and analyze the data, we normalize the number of cases in each age group (*P*_*age*_) by two different values. For comparison between ages groups, we normalize by the maximum number of cases in the relevant group (*max*(*P*_*age*_). For understanding a specific age group over time, we normalize by the total number of positive cases (*P*_*tot*_). During our study we defined two types of populations, one being the total population in a city (or country) (*N*_*age*%_) and one being the susceptible population (only people who have not been vaccinated or recovered from COVID-19) 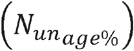. Note that we assume the immune population includes all recovered and vaccinated people and calculate the exact number using the equation in Table 1. We compare between *N*_*p*_ and the relevant population percentage (*N*_*age*%_ *or* 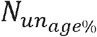) in order to estimate the change in positive cases over time in each age group. If this ratio is larger than 1 this indicates that the number of positive cases in the specific age group is larger than their proportion in the population. This shows which age groups are the main cause of the spread of the virus. In addition to the number of new positive cases, we focus on the number of accumulated cases in each city. Normalizing the accumulated cases by the maximum value allows for a better comparison between all cities regardless of their size (*A*_*c*%_). We also define two important factors which are used throughout the paper: *positive impact factor* and *hospitalization impact factor*. These factors are important to better understand the main age groups which effect reaching *community-immunity*. Cities in Israel differ due to many different parameters, we compare between cities with some similar parameters in order to reduce the effect of the parameters on our analysis. (for example, when analyzing the effect of median age, we compare cities with different social economic levels or cities that belong to different sectors).

**Table 1.** notations and definitions used throughout the paper

| Notation | Meaning | Origin/definition | Notation | Meaning | Origin/definition |
| --- | --- | --- | --- | --- | --- |
| $P_{age}$ | Daily positive cases in a specific age group | Corona Data | $H_{age}$ | Daily hospitalizations in a specific age group. | Corona Data |
| $P_{tot}$ | Daily positive cases in all age groups | Corona Data | $N_{tot}$ | Israeli population | CBS Data |
| $H_{tot}$ | Daily number of hospitalizations for all age groups | Corona Data | $N_{age}$ | number of individuals in a specific age group | CBS Data |
| $A_c(t)$ | Accumulated cases up to day t | Corona Data | $V_t$ | Number of individuals vaccinated first dose t week ago. | Corona Data |
| $N_{vac}$ | Number of immune due to vaccinations<sup>1</sup> | $0 V_0 + 0V_1 + 0.57V_2 + 0.66V_3 + 0.92V_{i>3}$ | $N_{imun}$ | Number of people who cannot be infected | $N_{vac} + N_{recovered}$ |
| $\hat{P}_{age}$ | Normalized number of cases | $\frac{P_{age}}{\max(P_{age})}$ | $P_{age\%}$ | Percentage of positive cases | $\frac{P_{age}}{P_{tot}}$ |
| $S_{tot}$ | Susceptible individuals | $N_{tot} - N_{imun}$ | $N_{age\%}$ | Percentage of the population | $\frac{N_{age}}{N_{tot}}$ |
| $N_{s\_age\%}$ | Percentage of Susceptible individuals | $\frac{N_{age}}{S_{tot}}$ | $\text{Impact}_{p-tot}$ | Positive Impact Factor (relative to $N_{tot}$) | $\frac{P_{age\%}}{N_{age\%}}$ |
| $\text{Impact}_{p-sus}$ | Impact of age group on positive cases (relative to $S_{tot}$) | $\frac{P_{age\%}}{N_{s\_age\%}}$ | $N_{adj\_imun}$ | Adjusted immune | $\text{Impact}_{p-tot} N_{imun}$ |
| $adj\_imun_{\%}$ | Percentage of adjusted immune | $\frac{N_{adj\_imun}}{N_{tot}}$ | $\text{Impact}_{H-tot}$ | Hospitalization Impact Factor | $\frac{H_{age\%}}{N_{age\%}}$ |

### Simulation methods

In order to simulate the spread of the pandemic in Israel and the vaccination policies we chose to use a stochastic implementation of an extended susceptible-exposed-infectious-removed (SEIR+) modeling framework^11^.

Unlike the deterministic classic SEIR model, this model uses an agent-based approach where each individual is represented as a node in a graph and has a state machine of possible states from the known SEIR model. In addition, there is a probability of transitioning between states which changes depending on the neighboring nodes in the graph.

The possible states are: S – susceptible, E – exposed, *I*_*pre*_ - Infected but pre symptomatic, *I*_*asy*_ – infected and asymptomatic, *I*_*sym*_ – infected and symptomatic, R – recovered, H – hospitalized, F – fatal (dead), *Q*_*s*_ – quarantined and susceptible, *Q*_*E*_ - quarantined and exposed, *Q*_*Pre*_ - quarantined and pre symptomatic, *Q*_*asym*_ - quarantined and asymptomatic, *Q*_*sym*_ - quarantined and symptomatic, *Q*_*R*_ - quarantined and recovered.

The transitions between each state are taken from known probabilities for COVID-19 aggregated from the Israeli data we used in this paper (exact numbers used can be found in our git repository). In addition, our model included testing campaigns, quarantines and vaccinations. We assume that each positive person puts the people in close contact to them in quarantine with a high probability.

Family members or any other individuals which are connected to the infected node go into quarantine and are tested. We also assume that a vaccinated person removes 80% of the interactions it has and therefore at a high probability does not spread the virus and cannot be infected.

The social graph is modeled to represent the Israeli population while taking into account family units, schools, workplaces and other social interactions. We separate all interactions into close contacts which occur on a daily basis (classmates, friends, housemates) and casual interactions which occur randomly between different people in the graph. We used the FARZ python package to model social interactions between different age groups. In addition, we assumed different family sizes based on the probabilities we found in the CBS data for Israeli family sizes and types. Eight different family structures were considered: single people, students, soldiers, couples without children (young and old), couples with children that are no longer living with them, couples with one young child, couples with one older child, couples with two children, couples with three children, couples with four children or more. We defined the probability of a family being in the graph and in addition within each family type we defined the probability of each family members age.

The nodes in the graph represent individuals, and the edges represent connections between individuals. The number of connections and their probability were fitted to match the known social connectivity matrix on the one hand^10^, and the pandemic behavior in Israel on the other. While connections within the community can be changed based on different government measurements, family connections remain the same throughout the pandemic and are broken only when a person enters isolation.

#### Statistical analysis

We chose to simulate a population of 15,000 nodes and all Fig. s from the simulation are normalized by the number of nodes for more clarity. We calibrated the model to match the *positive* and *hospitalization impact factor* and chose a graph that best matched Israel. These impact factors are not affected by vaccinations or lockdowns since they are the spread of the pandemic. The basis of our work here was that if the impact factors are similar to the real impact factors, the simulation demographics are similar to the Israeli one. We simulated vaccination policies with different vaccination rates, where each time we assumed the number of vaccinations per day is chosen from a normal distribution defining the mean number of vaccinations per day. (from 50 to 275). For each vaccination rate, we simulated the pandemic propagation without government intervention with four vaccination policies: (1) *young to old*: Prioritizing the younger population, (2) *old to young*: Prioritizing older population, (3) *triangle*: vaccinating the population over 60, then the younger population (16 to 35), then the remaining population, (4) *all ages*: vaccinations are equally distributed between all ages. To avoid stochastic influence, we repeated this process ten times with different seeds, and present the mean and variance of the results.

## RESULTS

### Notations and definitions

Throughout the paper we analyzed the data using different normalizations, notations and definitions. Table 1, defines all notations used.

### Overall pattern of the virus spread in Israel

Between March 2020 and January 2021, Israel faced three COVID-19 waves with a significantly higher number of cases in the second and the third waves. During each wave, following an increase in the number of new cases, the government introduced a lockdown to reduce the number of cases. One week into the third lockdown, Israel initialized its vaccination campaign.

As shown in Fig. 1a-b, the pattern of normalized positive cases in each age group 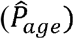 was similar during most of the pandemic - until the start of vaccinations. Notably, once the first selected population at risk began receiving vaccinations, there was an increase in new cases among the unvaccinated population (*transition phase*, Fig. 1b). When 48% of the population was either vaccinated or recovered, a significant decrease in new cases was observed across all age groups (*community-immunity phase*, Fig. 1b). Such a decrease in the number of new cases was not demonstrated following the second national lockdown, where the reopening of the economy resulted in an increase in the number of new cases (also reported by Rossman et al).^4^ We believe that this phenomenon is the beginning of the *community-immunity phase*, and will show in following graphs that the change between the two phases can be predicted and partially understood.

**Fig. 1.**
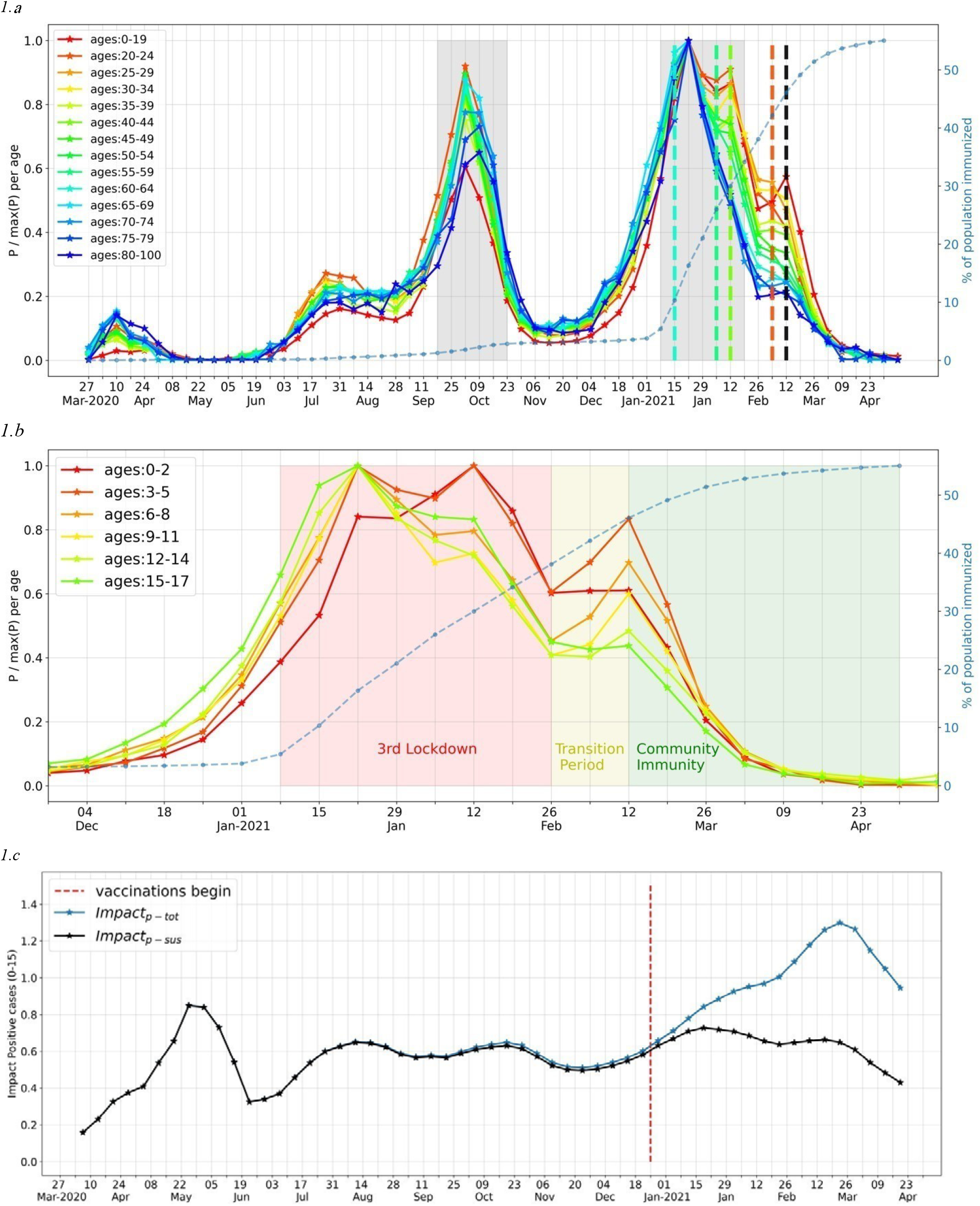
**a - Normalized positive cases by age 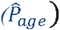 during the pandemi,** Values on the left y axis range between [0,1] in which higher numbers indicate more cases for that age group at time t. It seems that all ages reached their maximum value at the peak of the third wave (around 22.01.2021). The grey areas indicate lockdowns in which the government restricted social gatherings, including schools, offices and any other gathering with over 10 people. The right y axis and the doted blue line indicates the percentage of total immunized individuals (imun_%_). The colored dotted horizontal lines indicate the week in which individuals from the relevant age group started to be vaccinated (the colors match the colors of the legend). The black dotted line indicates the point at which cases started dropping for all age groups (regardless of their vaccination state) **b - Normalized positive cases during the 3rd wave for the unvaccinated young population** 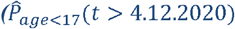. Population age was divided according to the policies in Israel while exiting the lockdown. During the transition period older children (≥ 13 years) did not go back to school and therefore there was no rise in cases indicated for their age group (15-17 years old). In addition, most infants, ages 0-2 years old were kept home and did not experience a rise in cases. As the immunized community reached 45%, the young population encountered a drop in cases despite the removal of all government restrictions. **c - Impact of unvaccinated age groups on positive cases relative to N_tot_ vs S_tot_**. This graph shows (marked by blue) the Impact of unvaccinated ages (0-15) on positive cases relative to the total population (impact_p-tot_ and (marked by black) the Impact of unvaccinated ages (0-15) on positive cases relative to the susceptible population (S_tot_) Impact_p-sus_. Values close to 1 indicate that the proportion of positive cases is the same as the ratio in the population. For this age group the ratio oscillates around 0.6, indicating that children in this age group had fewer positive cases than their proportion in the population. The red horizontal line indicates the beginning of the vaccination campaign in Israel.

#### The transition phase

To better understand the *transition phase*, we conducted an in-depth analysis of the young population. We calculated the *positive impact factor* of the young population relative to the entire population (Impact_*p*−*tot*_) and susceptible population (Impact_*p*−*sus*_) (Fig. 1c). Prior to the national vaccination campaign, (left of the dashed red line), Impact_*p*−*tot*_ was oscillating around 0.6, indicating that the young population’s positive ratio was 60% compared to its ratio in the total population. Once vaccinations started (right of the dashed red line), this ratio increased, reaching a maximum of 1.3, thus suggesting a “shift” in the disease – from the entire population to the unvaccinated people. This increase may be explained by normalizing the positive cases by the percentage of the susceptible population (Impact_*p*−*sus*_) rather than that of the whole population. Once normalizing by the susceptible population, the increase in the *positive impact factor* did not exist, indicating that the susceptible population is younger when vaccinating the older population.

#### The driver of community-immunity

To understand the effect of vaccinations on the spread of the disease and to examine the effect in different regions of Israel, representing different demographics, we examined the effect of vaccinations in 250 cities across Israel. After initially plotting the normalized number of cases 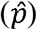 for all cities, we revealed a crucial effect for the median age of the city’s population in determining the accurate value of *community-immunity* (Fig. 2a): cities with a younger population presented a decrease in the number of new cases at an earlier percentage of immunized population with approximately 20% difference in the threshold among them. Interestingly, the entire country, which has a median age of 30.5 years, reached the same decrease in the number of new cases at 50% immunization (similar to Fig. 1a-b). This decrease was correlated with median age alone and not with the population sector (Modiin Ellit, an Ultra-orthodox Jewish city, and Ar’ara, an Israeli Arab city, reached the decrease at a similar percentage), or socio-economic level (Bat Yam, a city belonging to the lower socioeconomic rank compared to Qesariya, a city belonging to the upper socioeconomic rank).

**Fig. 2.**
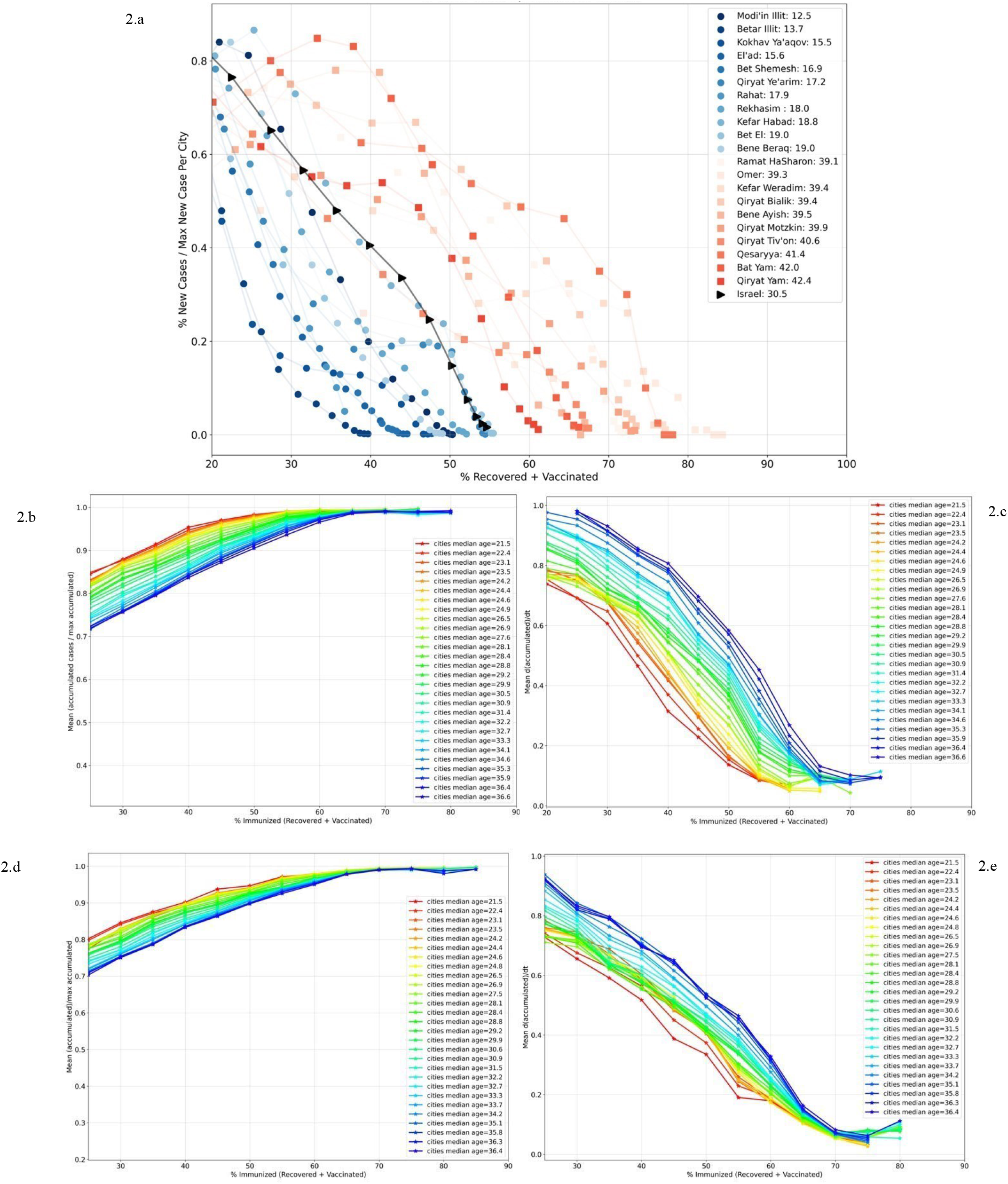
**a - The effect of vaccinations in cities across Israel.** This graph presents corona data from 20 cities that have the highest and lowest median ages in Israel (their median age is presented in the legend) In order to compare only the change in new case (eliminating differences in initial state), we plotted the number of new positive cases normalized by the maximum value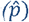. The X axis presents the percentage of immunized individuals in the relevant city (imun_%_). The black line shows the same relation for the entire country (median age of 30.5). **b - Normalized accumulated cases** 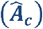 **as a function of immune individuals (imun_%_) and median age**. Each line represents the mean values of accumulated cases over 50 cities 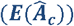. As the percentage of immunized population increased, the number of new cases decreased, reaching a plateau once 50-70% were immunized. In addition, the pace at which this plateau is reached changed according to the median age of the population. (More information in the methods section). **c - The derivative of normalized accumulated cases** 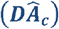 **as a function of immunized individuals (imun_%_) and median age**. This graph presents the central derivative of Fig. 2b. Lower values in this Fig. indicate a lower increase in new cases. Cities with a younger median age reached lower derivative values at lower percentages of immunized population. (more information in the methods section). **d - Normalized accumulated cases** 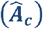 **as a function of adjusted immunized individuals (adj_imun_%_) and median age**. Similar to 2-b but here the X axis is the percentage of immunized people in which each vaccinated or recovered individual was multiplied by its age positive impact factor. As the percentage of adjusted immunized population increased, the number of new cases decreased, all reaching a plateau at approximately 60%. **e - The derivative of normalized accumulated cases** 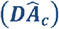 **as a function of adjusted immunized individuals (adj_imun_%_) and median age**. This graph is the derivative of Fig. 2d.

In addition, we evaluated the mean values of normalized accumulated cases in over 50 cities 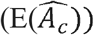 to raise the statistical confidence and lower the probability of other confounders. In order to achieve a large range of median ages we first sorted all cities by their median age, then pulled 50 cities at a time from the list with a stride of five from young to old cities (e.g. 0-50, 5-55, 10-60 etc.). For each batch of 50 cities we calculated 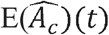, *E*(*N*_*imun*_)(*t*) and the median of the cities median age. The results presented in Fig. 2b-c match our previous observation (Fig. 2a), that a plateau occurs at higher percentages of immunization as the median age of the cities’ population increases (i.e., cities with younger populations reached the plateau at 50% immunization while cities with older populations reached it at 70%).

The results for Israel are different from the common belief that 60-75% of the population must be immune for herd immunity to be achieved (based on a theoretical R0 of 2.5-3).^5,6^ This percentage relies on the assumption that all individuals in the susceptible community have the same probability of spreading the virus. However, we show that this is not true for Israel and most likely for other countries. The real R0 for each individual is affected by a combination of demographic, social, epidemiological factors, as well as many other parameters. Understanding the exact contribution of each parameter is not possible; therefore, we chose to estimate the overall R0 for each individual based on their age.

To that end, we examined the distribution of new cases and COVID-19 related hospitalizations incidence for different age groups. As shown in Fig. 3a, the incidence of new cases (*P*_*age*%_) in ages 15-35 (blue shaded area) were higher than their proportion in the population (*N*_*age*%_), indicating that these ages are the spreading “engine” of the virus. However, the high number of cases in this age group was not reflected in elevated numbers of hospitalizations (*H*_*age*%_) which remained higher in elderly patients. To describe the ratio between the incidence of these parameters in each age group and the group’s proportion in the general population, we used the definitions *hospitalization impact factor* (Impact_*H-tot*_) and *positive impact factor* (Impact_*p*−*tot*_), respectively (Fig. 3b). The red shaded area shows that the ages over 60 experienced more hospitalizations despite having a low *positive impact factor*. The *hospitalization impact factor* is important for avoiding the overload of hospitals and the *positive impact factor* can describe the effect of vaccinations on the spread of the disease across different ages. The findings of this analysis highlight the differences between age groups in terms of their contribution to both the transmission of the disease, and their contribution to the hospitalizations. Realizing that each age group contributes differently to the spread of the pandemic highlighted the need to treat each vaccinated person in each city as an individual rather than assuming that everyone is an equal contributor to reaching *community-immunity*. We therefore chose to multiply each vaccinated or recovered person by the *positive impact factor* relevant for their age group. We demonstrated this hypothesis by implementing this technique on the data in Fig. 2b-c. Fig. 2d-e shows that the different median ages collapse to almost the same value of 60% immunizations needed for *community-immunity*. These findings demonstrate that most differences among the various cities were caused by the *positive impact factor* of each vaccinated person, implying that the observed difference among cities is mainly due to their different age structure.

**Fig. 3.**
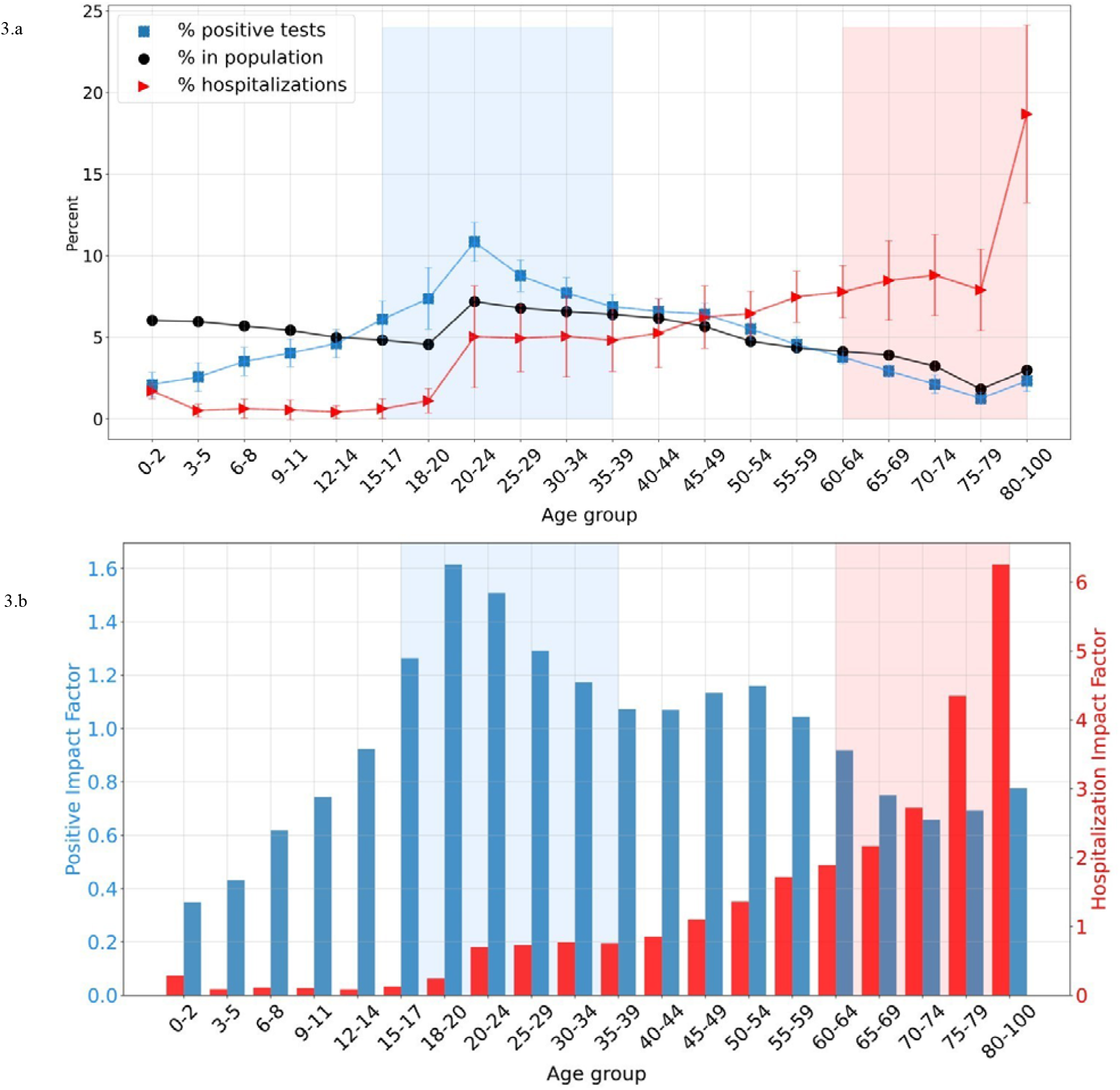
**a - Percentage of positive cases** P_age%_ (blue), **hospitalizations** H_age%_ (red) **and population** N_age%_ **(black) in each age group.** Each line indicates the number of positive cases, hospitalizations and people in the relevant age group normalized by the total number of positive cases, hospitalizations and population. This Fig. shows the contribution of each age group to the spread of the pandemic (marked by blue) or to the hospitalizations (marked by red). When the blue/red line are above the black we assume these age groups contributed more to the spread of the pandemic/hospitalizations respectively. The blue shaded area depicts age groups that have a high proportion of positive cases (higher than their proportion in the population). The red shaded area shows age groups that have a high proportion of hospitalizations. **b - Percentage of positive cases normalized by percentage of the population (blue), and percentage of hospitalizations normalized by percentage of the population (red)**. This Fig. is a continuation of Fig. 3a. The blue columns represent the blue line divided by the black one in Fig. 3a. Values above 1 show that the percentage of positive cases/hospitalizations was greater than their percentage in the population. Two important age groups are noted: those responsible for spreading the virus (blue shaded area) and those responsible for occupying the hospitals (red shaded areas).

#### Vaccination policy

In order to better understand the effect of the vaccination campaigns and rate on the overall positive cases and hospitalizations we compared different vaccination policies in a simulated environment. The *positive* and *hospitalization impact factor* present two opposing forces when trying to evaluate the best vaccination policy. On the one hand, vaccinating the age groups with the highest *positive impact factors* will reach *community-immunity* faster, on the other hand, vaccinating based on the *hospitalization factors* will result in immediate relief for the hospitals’ burden. We simulated the pandemic without government intervention using a stochastic implementation of an extended susceptible-exposed-infectious-removed (SEIR+) modeling framework^11^. We formulized Israel’s contact network in a graph where links in the graph are based on family cells, social gatherings and community connections. We compare five different vaccination rates with four vaccination policies: (1) *young to old*: Prioritizing the younger population, (2) *old to young*: Prioritizing older population, (3) *triangle*: vaccinating the population over 60, then the younger population (16 to 35), then the remaining population, (4) *all ages*: vaccinations are equally distributed between all ages. To avoid stochastic influence, we repeated this process ten times with different seeds, and present the mean and variance of the results.

The final accumulated cases (after reaching *community-immunity*) of the four different policies are presented in Fig. 4a. As expected, prioritizing age groups with high *positive impact factors* results in a lower number of accumulated cases regardless of vaccination rate. The effect however, seems to degrade from 50% for fast vaccination rates to 10% for slow vaccination campaign.

**Fig. 4.**
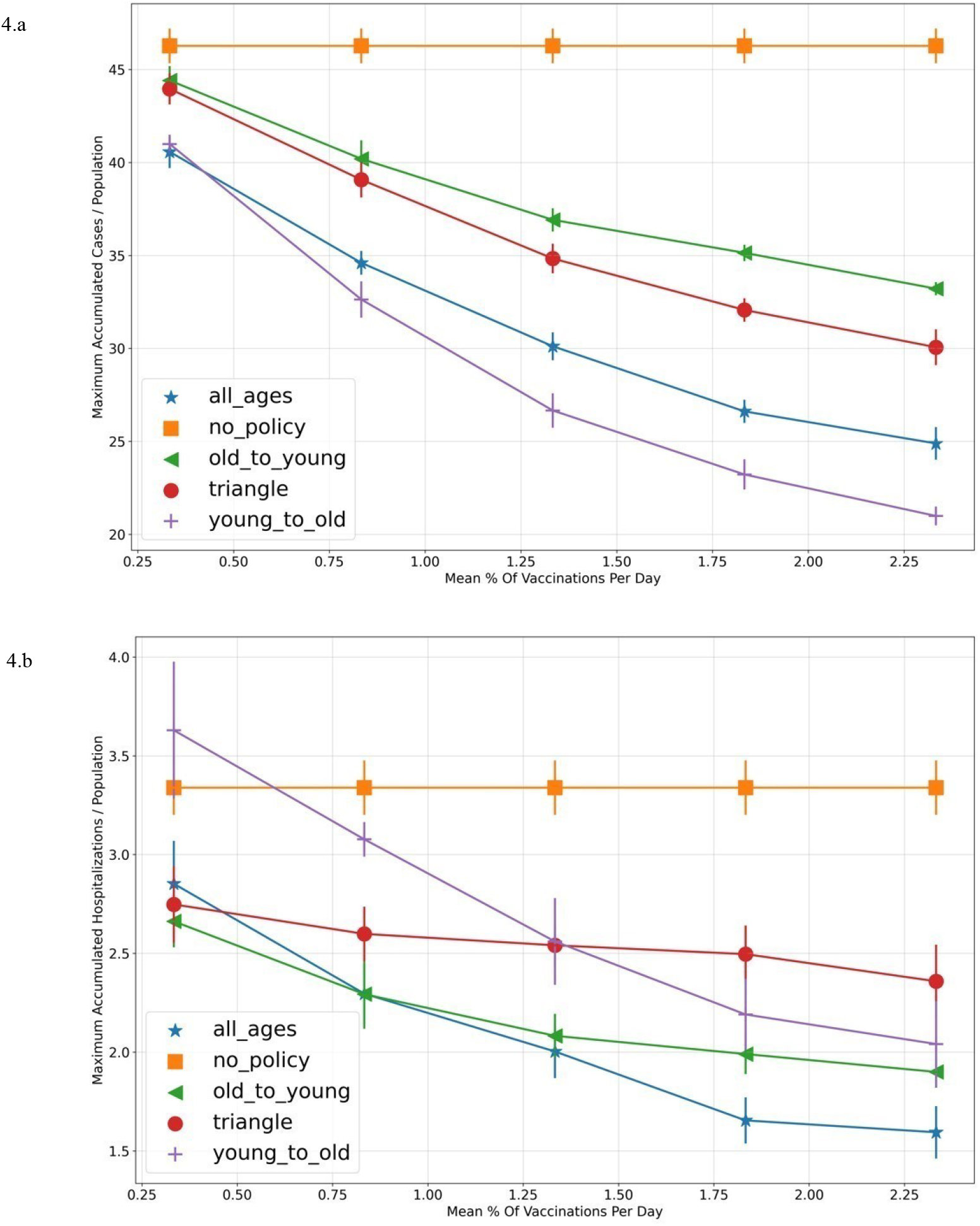
***a* - Final accumulated cases % (after reaching community-immunity) as a function of vaccination rate and vaccination policy. *b -* Final hospitalization cases % (after reaching community-immunity) as a function of vaccination rate.** Each point is a mean and variance of ten seeds for a specific vaccination policy and vaccination pace. The X axis indicates the mean percentage of vaccinations per day (mean number of vaccinations per day divided by the population size). The different policies are: (1) “young to old” :Prioritizing the younger population (vaccinating based on age from the youngest people to the older ones), (2) “old to young” :Prioritizing older population (vaccinating by age from the oldest population to the youngest), (3) “triangle” :First prioritizing age groups with high hospitalization impact factor (Fig. 4b marked by red) then prioritizing age groups with high positive impact factor (Fig. 4b marked by blue) and then the rest (vaccinating the population over 60, then vaccinating the younger population from 16 to 35, then the rest of the population by age), (4) “all ages” : No Prioritizing – vaccinations are equally distributed between the age groups.

The *hospitalization factor* (Fig. 4b), on the other hand, is much less predictive, as vaccinations affect the non-vaccinated population and change the probability of hospitalizations in the population. Fig. 4b presents two interesting results; Firstly, it seems that not prioritizing any age group is in most cases the best policy for reducing hospitalizations. Secondly, prioritizing the younger population might result in more hospitalizations than not vaccinating at all (when the number of vaccinations per day is low). This surprising result can be due to the *transition phase* described above which creates a “shift” in the disease spreading– from the entire population to the unvaccinated population. In this case, prioritizing the young population shifts the spreading to the elderly, which are more likely to be hospitalized. Therefore, although the number of positive cases is reduced, the probability of each positive case being hospitalized increases as the susceptible population is older.

Our simulation shows that prioritization and vaccination rate have an immense effect on the overall hospitalizations and number of cases. In Fig. 4-b, we show a difference of almost 40% between vaccination policies which maintains regardless to the vaccination rate. Despite the common belief that vaccinating the older population first will lead to a successful campaign, our model shows for most vaccination rates, that vaccinating everyone at the same pace (*all ages*) will achieve the best outcome. This strategy seems to work better than all other methods since it reduces the spreading population (by vaccinating people from the younger community) on the one hand, while lowering the number of potential hospitalizations (by vaccinating the older population) on the other. We note that our simulation is based on Israel’s impact factors and the graph represents the Israeli population, but the simulation is general and can be adapted to other countries easily.

## DISCUSSION

In this study, we examined the effect of the COVID-19 vaccination campaign on 250 cities in Israel each with unique demographic characteristics and ethnic groups. Our analysis revealed that the median age of the city’s population has a crucial effect in determining the accurate value of *community-immunity*: cities with younger populations reached a decrease in new cases when a lower percentage of their residents were immunized with approximately 20% difference in the threshold between these cities compared to older ones. These findings indicate that cultural similarities or compliance to government restrictions may have less significance in light of an effective vaccination program.

Our findings corroborate those of a previous analysis conducted in Israel approximately 2 months after the start of the vaccination campaign, which showed that the decrease in the clinical measures occurred only after more than 50% of the population in a given age group had been vaccinated by the first dose or recovered and that the effect was greater in cities and geographical statistical areas in which a higher fraction of individuals were vaccinated earlier.^4^ To understand the effect of the vaccinations on the pandemic, we developed two new factors: the *hospitalization impact factor* and the *positive impact factor*. The *hospitalization factor* is important for avoiding the burden on hospitals and the *positive impact factor* for containing the virus and minimizing its spread.

By adjusting the percentage of recovered and vaccinated individuals in the population based on the *positive impact factor* for each individual and redefining the immunized population, we demonstrated that most differences among cities were originated by the *positive impact factor* of each person. These findings not only affect our understanding of *community-immunity* but might also help estimating the timing in which other cities, that have only started vaccinating, will reach *community-immunity*. Based on these impact factors, it is clear that vaccinating older communities is necessary for relieving the burden on hospitals while vaccinating younger adults is needed for containing the spread of the virus and reaching *community-immunity*. The higher impact factors in the young population may be attributed to many interactions and a relatively low risk for this group, which may contribute to less careful behavior and higher numbers of infections amongst them.^7-9^

The common estimate is that 60-75% of the population must be immunized to achieve herd immunity.^5,6^ This might contradict what we observed in Israel, where *community-immunity* was achieved when the percentage of immune individuals was approximately 50%. However, adjusting the data to the *positive impact factor*, showed that all cities in Israel achieved *community-immunity* at 60-70%. This is well explained by the R0 assumption of equal probability for spreading the virus. Therefore, adjusting for a combination of demographic, social, epidemiological and many other parameters reflects the real R0 for each individual. Finally, we evaluated four different vaccination policies (*young to old, old to young, triangle* and *all ages*) using a stochastic implementation of the extended seir model. As expected, the final number of cases is lower when prioritizing using the *positive impact factor*, i.e. younger population first and older population later. However, the final number of hospitalizations is more complex and depends on the vaccination rate. In general, it seems that not prioritizing ages results in less hospitalizations while *young to old* policy might even result in a larger number of hospitalizations compared to not vaccinating (due to the *transition period*). Taking both factors into account, we show that vaccinating all ages simultaneously achieves a better outcome compared to other policies.

In conclusion, our findings show the impact of the countries age structure on the success of the vaccination campaign, and the need to redefine the immunized population which leads to *community-immunity*. We believe our analysis can assist other countries to better predict the outcome of vaccinations on their COVID-19 cases and also improve their vaccination policy.

## Data Availability

The data included two main COVID-19 tables given to us by the MoH. The first with individual identification number (random number for ethical and anonymous reasons), age group, geographic region, date of polymerase chain reaction (PCR) test for identification of COVID-19 and test result, time of hospitalization, worst case while hospitalized, COVID-19 outcome (alive/dead). The second table included vaccination data, with data about the number of vaccinations per day for different ages and geographic regions. Large portions of the aggregated data used in this work is publicly available in Hebrew divided into 6 different databases. Sensitive information, mostly related to the hospitalization status is not publicly accessible. In addition to the COVID-19 data we used data from the CBS (Central Bureau of Statistics) regarding cities median age, number of people in each age group and data about families in Israel. The COVID data included 262 cities with over 5000 citizens, but only 250 were taken into account since 12 cities did not have information on their median age
Our analysis code in addition to some of the publicly available data, can be found in this Git repository

https://github.com/CoronaPolicy/prediction_model

## Acknowledgment

This work was supported by the Bosch Center for Artificial Intelligence (BCAI). The Bosch Group as a whole has been involved in the fight against the global coronavirus pandemic from the very beginning.

This work was also supported by Assuta Medical Centers.

The authors would like to thank the Israel Ministry of Health for assisting in obtaining the raw data. We would also like to acknowledge Mr. Medad Hoze and Mr. Yovav Sanders for their contribution to the virtual data-challenge event. Last, we would like to thank Dotan Di Castro and Nuria Oliver for their guidance and support while doing this work.

## Author contribution

C.R. and O.S. conceived the project, designed and conducted the analyses, interpreted the results, wrote the manuscript and revised it critically for important intellectual content. M.T. made substantial contributions to conception and design and acquisition of data. Y.W has been involved in drafting the manuscript and revising it critically for important intellectual content and R.B made substantial contributions to conception and design, has been involved in writing the manuscript and revising it critically for important intellectual content.

## Appendix

### A. Simulation state graph

Our model (Fig. S2) extends the classic SEIR model by further dividing the infectious subpopulation into pre-symptomatic (Ipre), asymptomatic (Iasym), symptomatic (Isym), and hospitalized (severely symptomatic, IH. In addition, this model also accounts for isolation-based interventions (e.g., isolating individuals in response to testing or contact tracing).

### B. Validating our social connection graph

The social graph is modeled to represent the Israeli population while considering family units, schools, workplaces and other social interactions. In order to validate our choices, we calculated the positive and hospitalization impact factor during the spread of the virus in our simulation. The results presented in Fig. S3 match our observations during the pandemic in Israel [figure 3-a main text].

**Fig. S2.**
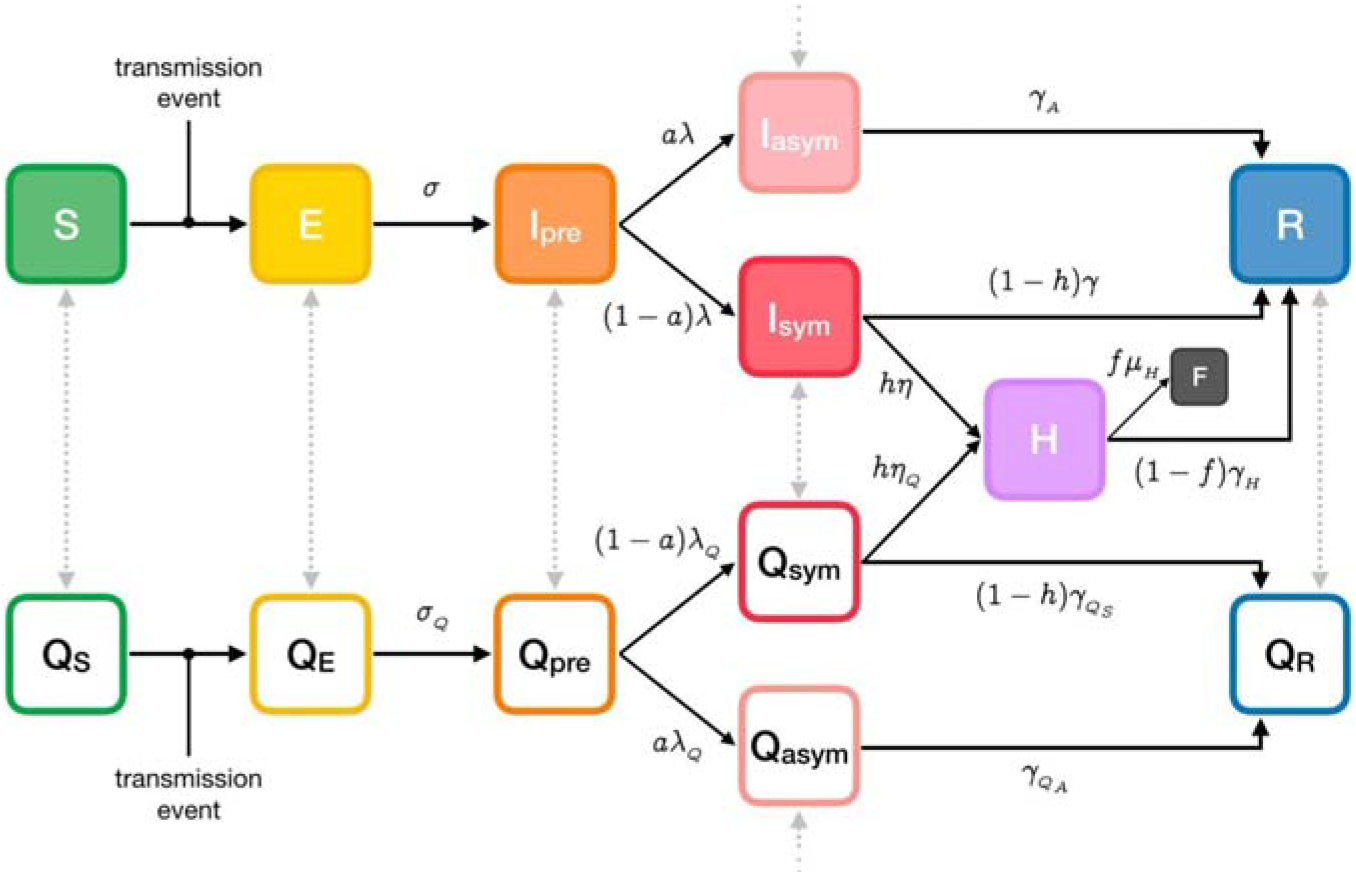
seir+ model state machine

**Fig. S3.**
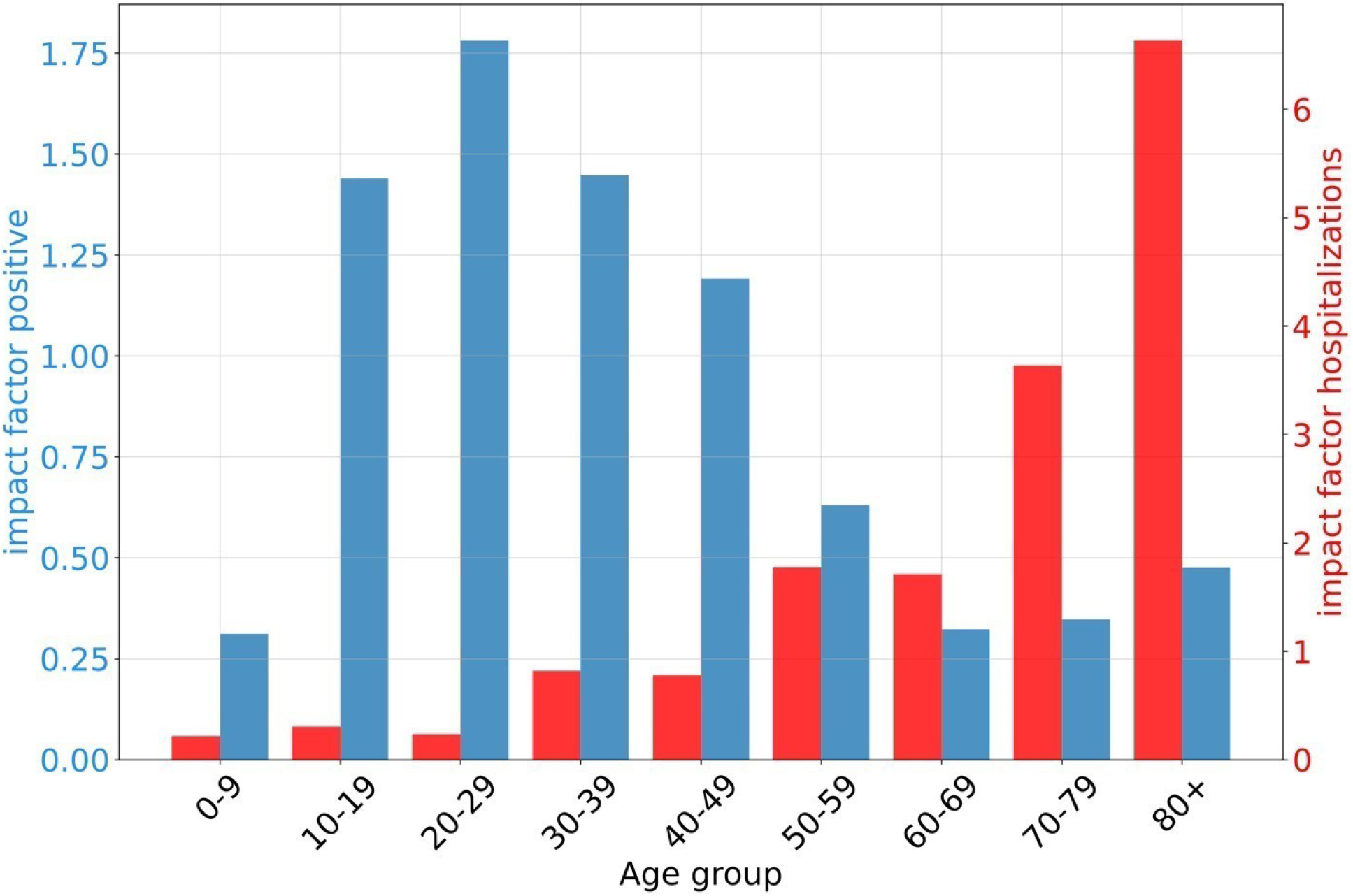
positive and hospitalization impact factors from the simulation for each age group

